# Axenfeld-Rieger syndrome associated with a megabase-scale inversion separating *PITX2* from a conserved enhancer locus

**DOI:** 10.1101/2025.06.05.25327661

**Authors:** Lucas A. Mitchell, Joshua Schmidt, Emmanuelle Souzeau, Lachlan S. W. Knight, Giorgina Maxwell, Andrew Dubowsky, Ridia Lim, Edward Formaini, Matthew Welland, Cas Simons, Daniel G. MacArthur, Janey L. Wiggs, Jamie E. Craig, Owen M. Siggs

**Affiliations:** Genomics and Inherited Disease Program, Garvan Institute of Medical Research, Sydney, New South Wales, Australia; Faculty of Medicine, University of New South Wales, Sydney, New South Wales, Australia; Department of Ophthalmology, Flinders University, Adelaide, Australia; SA Pathology, Adelaide, Australia; Sydney Eye Hospital, Sydney, New South Wales, Australia; Centre for Population Genomics, Garvan Institute of Medical Research and UNSW Sydney, 384 Victoria Street, Sydney, Australia; Centre for Population Genomics, Murdoch Children’s Research Institute, Melbourne, Australia; Department of Ophthalmology, Harvard Medical School, 243 Charles Street, Boston, MA 02114, United States

## Abstract

Axenfeld–Rieger Syndrome (ARS) is an autosomal dominant condition with both ocular and non-ocular manifestations. ARS is primarily caused by coding variants at the *PITX2* or *FOXC1* loci, yet many cases still remain undiagnosed. Here we used whole-genome sequencing to identify two non-coding structural variants associated with a typical presentation of *PITX2*-associated ARS: one with a 450 kb deletion removing a series of conserved enhancer elements distal to *PITX2*, and the second with a 12.5 Mb inversion displacing the *PITX2* gene from these same enhancer elements. Neither variant disrupted the *PITX2* gene itself, and therefore both were expected to reduce *PITX2* expression by disrupting its proximity or access to enhancer elements. Enhancer-disrupting intergenic inversions therefore represent a unique genetic mechanism for the development of ARS, which should be carefully considered in the context of ARS and other conditions without a conclusive genetic diagnosis.

## Introduction

Axenfeld–Rieger Syndrome (ARS) is a rare autosomal dominant condition, with primarily ocular manifestations. Key ocular features of ARS include posterior embryotoxon, iridocorneal adhesions, corectopia, and polycoria, with up to a 75% increased risk of developing glaucoma.^1^ Non-ocular syndromic features include dental anomalies (hypodontia, microdontia), mild craniofacial dysmorphism, and umbilical anomalies (redundant periumbilical skin, umbilical hernia).^2–4^

ARS, as well as glaucoma associated with non-acquired ocular anomalies, is primarily caused by variation in *PITX2* or *FOXC1*.^5^ Variants in these two genes account for approximately 70% of ARS cases, although it is unclear whether the remaining 30% are caused by undetected variants at these same loci, or others.^6^ Furthermore, the presence of microdontia/hypodontia and umbilical anomalies is highly specific for *PITX2*-associated ARS: in one case series these were observed in 91% and 94% of cases with *PITX2*-associated ARS (n=59), as compared to 0% and 11% of *FOXC1*-associated ARS (n=69).^6^

ARS-associated variants in *PITX2* have provided some important insight into this question. Initially reported in 1996, *PITX2* encodes a bicoid-like homeobox transcription factor that plays a vital role in regulating transcription during embryogenesis, particularly in the development of the eye and its anterior segment.^7^ The locus encodes at least three isoforms, one of which (*PITX2*c) exhibits cardiomyocyte specificity, and putative gain-of-function missense variants unique to this isoform (p.Pro41Ser) have recently been associated with atrial fibrillation.^8^

Most ARS-associated *PITX2* variants directly disrupt the canonical open reading frame via single nucleotide variants and short indels, or partial and full-length gene deletions.^6^ However, upstream non-coding deletions of different sizes have also been described in at least seven unrelated ARS families.^9–14^ These deletions all disrupt a series of conserved essential regulatory elements, with a common critical interval spanning three such elements (CE5-7, hg38 chr4:110926950-111115957).^9,14^ Another two studies have identified balanced inversions or rearrangements disrupting *PITX2*.^15,16^ These variants all highlight the critical importance of conserved enhancer loci in ARS, as well as the need to carefully assess structural variation when investigating the genetic basis of ARS.

Here we describe a unique genetic mechanism of ARS involving disruption of the *PITX2* distal enhancer locus. Through short-read whole genome sequencing (WGS) and structural variant calling, we identified two non-coding structural variants: the first a non-coding 450 kb deletion, and the second a 12.5Mb inversion segregating within a family with a typical *PITX2*-associated ARS phenotype. While the *PITX2* gene itself was not disrupted by either variant, its proximity to conserved enhancer loci was, with the inversion causing a significant spatial displacement of the gene from its enhancer.

## Methods

### Study participants

Patients and family members were recruited as part of the Australian and New Zealand Registry of Advanced Glaucoma.^5^ Written informed consent was provided under protocols approved by the Southern Adelaide Clinical Human Research Ethics Committee (305–08), and adhering to the tenets of the revised Declaration of Helsinki.

### Exome and genome sequencing and analysis

DNA was prepared from venous blood samples, after temporary storage at -80°C, using the QIAGEN DNeasy Blood and Tissue Kit (Hilden, Germany) and according to the manufacturer’s instructions. Whole exome sequencing was performed as previously described.^17^ PCR-free WGS was performed by the Clinical Research Sequencing Platform at the Broad Institute. Library construction was performed using a KAPA HyperPrep kit, with libraries dual-barcoded and sequenced with 150 bp paired-end reads to a mean coverage of 30x. Alignment and calling of single nucleotide variants (SNVs) and insertions/deletions (indels) was performed using the Illumina DRAGEN (Dynamic Read Analysis for GENomics) pipeline. Structural variants (SV) were called using the GATK-SV pipeline (https://github.com/broadinstitute/gatk-sv).^18^ All variants were analysed using the *seqr* platform, and are represented in hg38 coordinates.^19^

### SNP genotyping and CNV calling

Individuals from Family 1 were genotyped on either HumanOmniExpress-12-v1-0-K (I:2) or HumanCoreExome-24v1-0_A genotype (II:2) arrays. Each of the individuals reported here were members of larger batches of samples, with each batch assessed for quality using Illumina GenomeStudio. After export of ‘Genotype’, ‘Log R Ratio’ and ‘B Allele Frequency’, samples were assayed for CNV using *acne* (available at https://github.com/joshuamschmidt/acne), a nextflow pipeline for CNV calling that utilises PennCNV^20^ as the CNV caller. CNV calls were annotated and filtered using thresholds recommended in the PennCNV documentation.

## Results

We investigated two kindreds both with a combination of ocular and systemic features typical of *PITX2*-associated ARS (Table 1), and phenotypic segregation suggesting autosomal dominant inheritance.

**Table 1:**
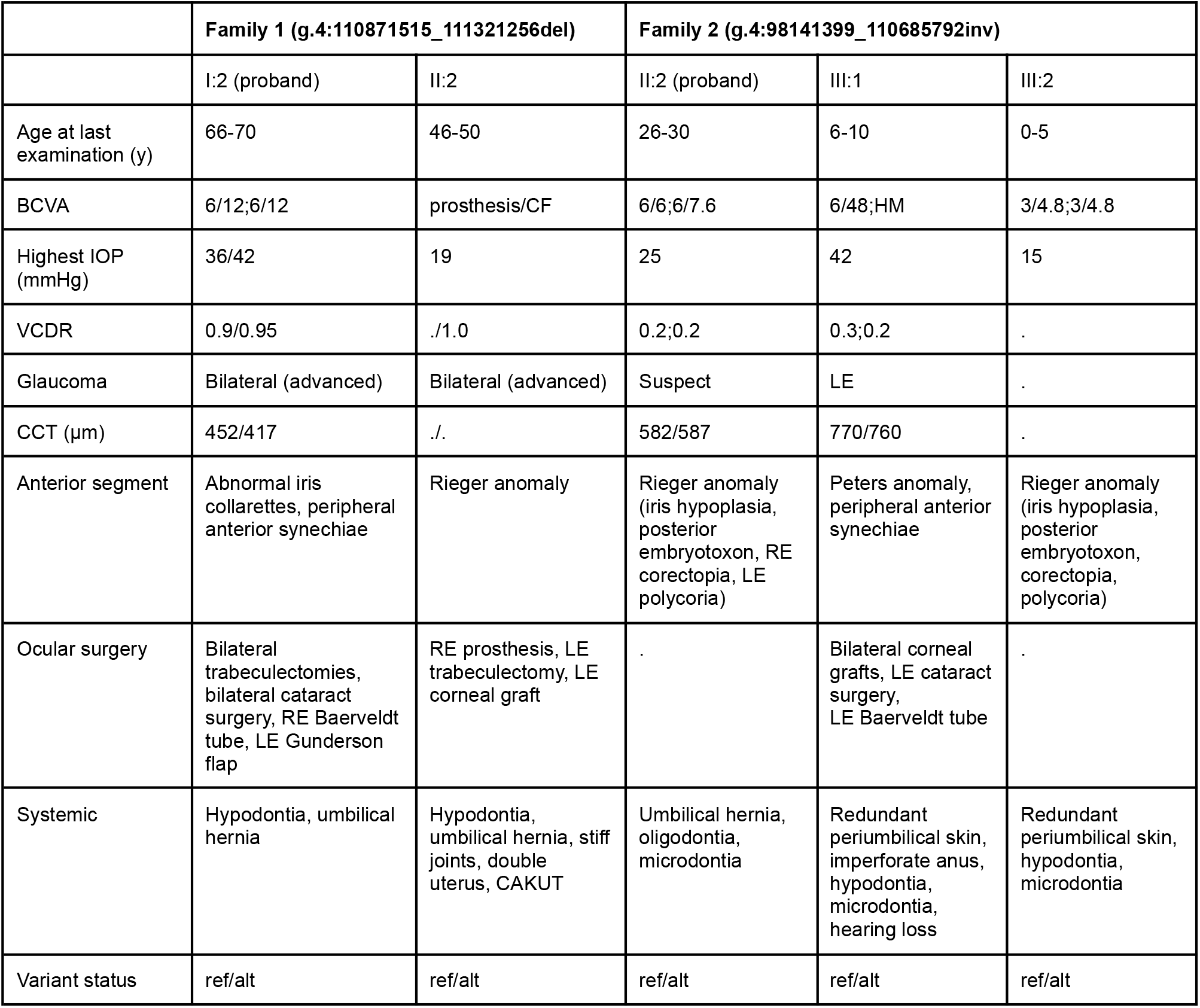
Clinical features of recruited family members. Y, years; BCVA, best corrected visual acuity; IOP, intraocular pressure; VCDR, vertical cup-to-disc ratio; CCT, central corneal thickness; RE, right eye; LE, left eye; CF, count fingers; HM, hand movements, CAKUT, congenital anomalies of the kidney and urinary tract. Data is presented as RE/LE for each individual. Clinical data from individual III:1 (Family 1) was not available.

Family 1 was initially identified during a copy number variant analysis of genotyping array data (see methods), which revealed a potential non-coding deletion distal to the *PITX2* locus, inferred to be 482 kb in I:2 (g.4:110890464_111372690del) and 477 kb in II:2 (g.4:110891306_111368288del). These were further resolved on WGS, which confirmed a shared 450 kb deletion at (g.4:110871515_111321256del), overlapping a series of 6 conserved non-coding elements (CE5-9 and CE15, Figure 2B).

**Figure 1:**
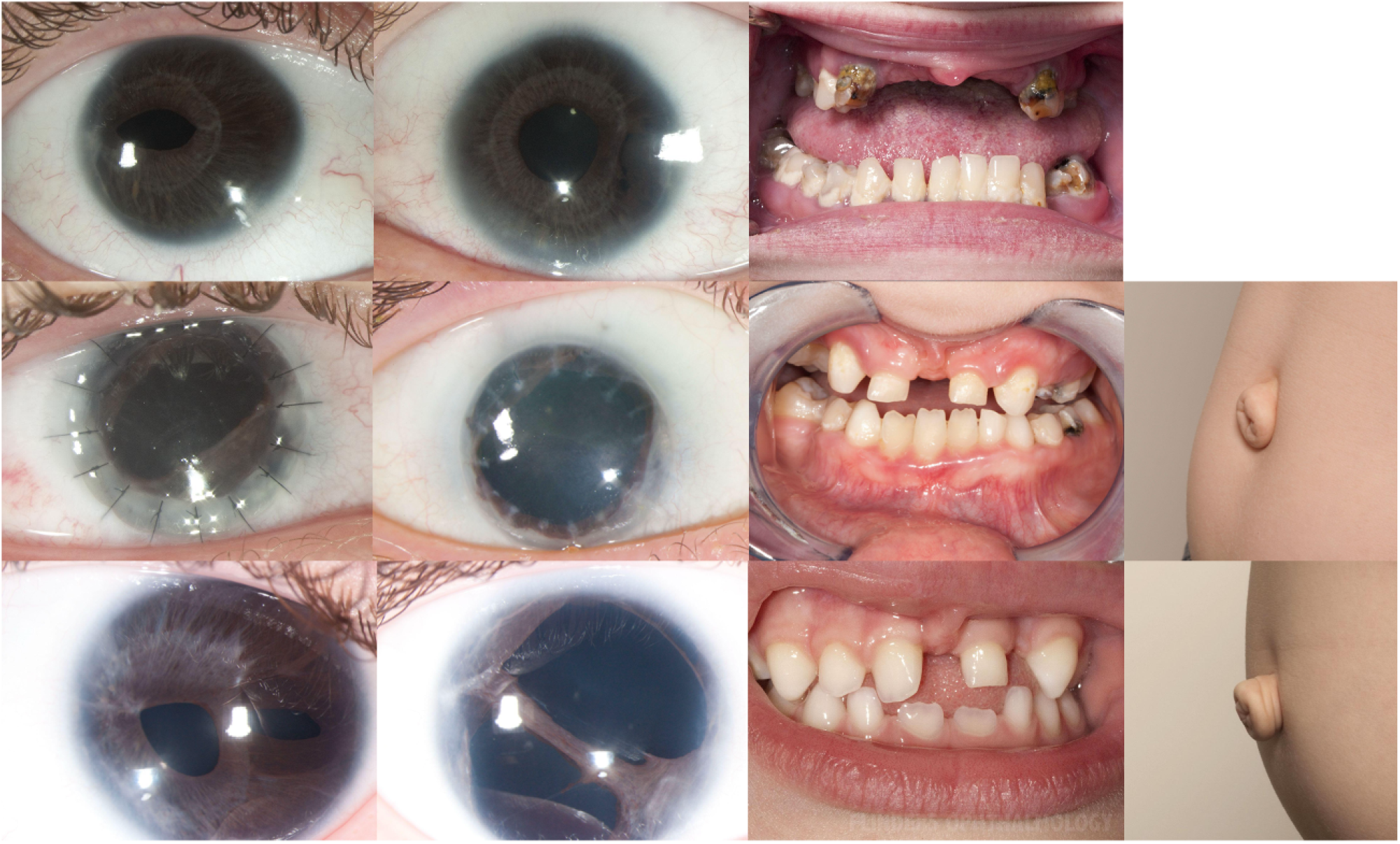
Ocular and systemic features of Axenfeld Rieger Syndrome. Photographs of all affected family members, including (from left to right) the anterior segment of the right eye, left eye, teeth, and umbilicus. Corectopia and dental hypoplasia are evident in all individuals, with redundant periumbilical skin documented in two individuals, polycoria in two individuals, and bilateral penetrating keratoplasties in one individual.

**Figure 2:**
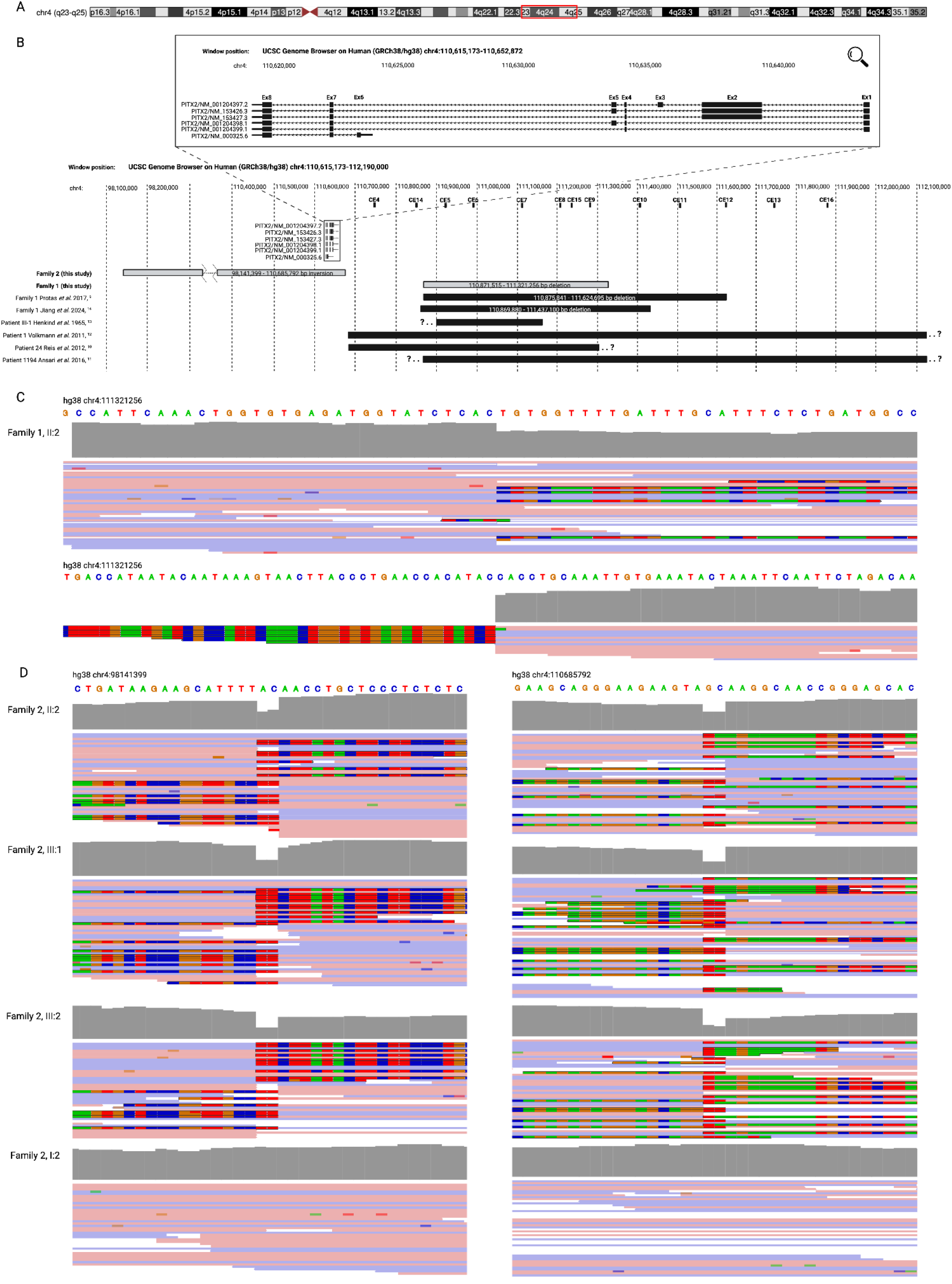
A 12.54 Mb inversion and 450 kb deletion at the *PITX2* locus. (A) Schematic of chromosome 4, highlighting the g.4:98141399_110685792inv region in red.^21^ (B) Schematic of the *PITX2* locus and surrounding region, demonstrating the location of the deletion and inversion variants described here (hg38, g.4:110871515_111321256del and g.4:98141399_110685792inv), along with previously reported *PITX2* enhancer deletions. Adapted from Protas et al. 2017.^9^ Note that only the *PITX2* gene is shown for simplicity: another 65 genes are within the inverted region indicated for Family 1. Read-level support for the g.4:110871515_111321256del (C) and g.4:98141399_110685792inv (D) variants. The absence of mapped reads at the proximal breakpoint in (C) is due to the presence of a common 4.89kb deletion *in trans* (4:111316365-111321256, gnomAD v4.1.0 NFE AF = 0.1243).^22^

Based on the clinical presentation of Family 2 (II:2), in 2013 the complete coding sequence and intron-exon boundaries of *FOXC1* and *PITX2* were amplified by PCR and covered by capillary sequencing, and analysed for copy number variation by multiplex ligation-dependent probe amplification (MLPA), although neither assay revealed a likely genetic cause. Individual II:2 was also referred for karyotype studies and array CGH (BlueGnome Cytochip oligo ISCA (4080-5) 8x60K array, 0.2Mb resolution), including a detailed examination of the *FOXC1* and *PITX2* loci. Both assays were reported as normal.

Four members of Family 2 (I:2, II:2, III:1, and III:2) were then subjected to whole exome sequencing. There were no rare and predicted deleterious SNVs or short indels in any genes associated with anterior segment dysgenesis, including missense, nonsense, and essential splice variants, although non-coding and structural variants could not be systematically examined. Manual inspection of all coding exons of *FOXC1* and *PITX2* also did not reveal any significant copy number variants to suggest a deletion or duplication, nor did a genome-wide analysis of copy number variation based on exome or genotyping array data (Schmidt et al., in preparation).

Finally, the same four individuals were subjected to research-based whole genome sequencing. Again, close inspection of the *FOXC1* and *PITX2* loci did not reveal any rare SNVs or short indels with a predicted deleterious effect, nor did a careful manual inspection of coverage depth across both genes. A genome-wide structural variant callset was generated using the GATK-SV pipeline ^18^: although this did not reveal any suspicious insertions or deletions at the *FOXC1* and *PITX2* loci, it did reveal a 12.54 Mb inversion on chromosome 4 (hg38, g.4:98141399_110685792inv), encompassing 66 genes including all coding exons of *PITX2* (Figure 2). This inversion variant was present in a heterozygous state in the proband (II:2) and their affected children (III:1, III:2), but absent in II:2’s parent (I:2) who was unaffected. It was absent from the gnomAD SV v4.1.0 callset, and from a jointly-called dataset of 8920 alleles from a variety of rare disease patients and their family members.

Since *PITX2* was the most distal gene in the inverted region and adjacent to the distal breakpoint (TSS 43.7 kb proximal to the breakpoint) (Figure 2B), we hypothesised that displacement from an enhancer locus could explain the observed phenotype. This distal breakpoint was found to be proximal to a series of conserved *PITX2* enhancer loci ^9,14^, including those deleted in Family 1 and other previously reported families (Figure 2B). The inversion variant would then be predicted to displace (and invert) the *PITX2* gene another 12.54 Mb proximal to these enhancers, leading to a predicted reduction in *PITX2* expression and haploinsufficient effect.

## Discussion

Here we describe two ARS families with non-coding structural variants: the first a 450 kb deletion (g.4:110871515_111321256del), and the second a 12.54Mb inversion (g.4:98141399_110685792inv). Both disrupt a series of conserved enhancer elements distal to the *PITX2* gene, three of which (CE5-7, hg38 chr4:110926950_111115957) form part of a shared critical interval across 7 previously reported families with enhancer deletions (Figure 2B). In the case of the deletion observed in Family 1 these same elements are lost (in addition to CE8, CE9, and CE15), and in the case of the inversion in Family 2 these same elements become displaced from the *PITX2* gene body.

In light of recurrent non-coding deletions at the *PITX2* locus, the haploinsufficient nature of *PITX2*-associated ARS, and the highly specific phenotype observed here, the most likely consequence of the Family 2 inversion appears to be a loss of *PITX2* expression due to the displacement of the *PITX2* gene body from its enhancer loci. A similar mechanism may also be at play in two ARS cases associated with balanced translocations t(4;16) and t(4;11)), where the breakpoints lie near the *PITX2* gene although were not known to disrupt it.^23^ We also note that a similar gene-sparing inversion mechanism (involving the *PAX6* and *PITX2* loci) has also been mentioned in conference proceedings ^24^, although these are yet to be published.

In summary, in addition to enhancer-disrupting deletions, intergenic inversions represent a unique genetic mechanism for the development of ARS, which should be carefully considered in any ARS families, or indeed any suspected monogenic condition, where a causal variant has not been confidently assigned.

## Data Availability

All data produced in the present study are available upon reasonable request to the authors.

## Acknowledgements

Supported by the NIH (R01 EY031820-01 to JW, JEC, OMS), NHMRC (to LM, ES, JEC), and the Snow Medical Research Foundation (Grant No. PF2019-040 to OMS). The funders were not involved in the design of the study, collection, analysis, and interpretation of the data, the writing of this report, or the decision to submit the article for publication. We thank the patients and their families for their participation, Angela Chappell for ophthalmic photography, and Dr John Pater for clinical support. Figure 1A and Figure 2B were created using BioRender.com.

